## Supplemental tables and figures for "Identification of novel vertebral development factors through UK Biobank driven genetic and body imaging analysis reveals markers for back pain"

**This PDF file includes:**

Figs. S1 to S7
Tables S1 to S1

**Fig. S1.**

Associations between vertebral anomalies and physical measures including the standing height, sitting height, seated height, waist, and hip circumferences, FEV and FVC.

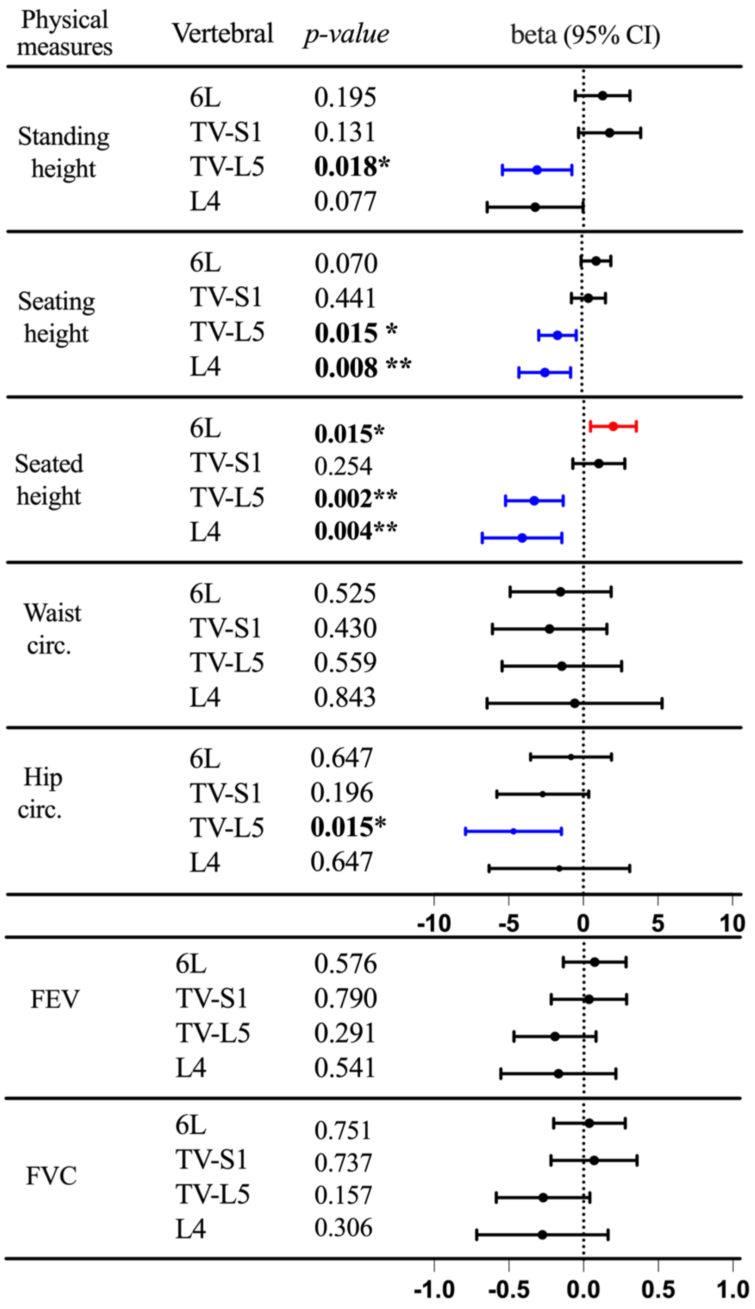

**Fig. S2.**

Associations between rib anomalies and the experience of pain lasting 3+ months (including headaches, neck/shoulder pain, and back pain). Odds ratio (OR) are calculated based on logistic regression models with sex and age as covariates. FDR adjusted one-sided *p*-values are displayed. Abbreviations: L1R: extra ribs on L1; 12R-hypo: hypoplastic 12$^{th}$ ribs; 11R: 11 rib pairs.

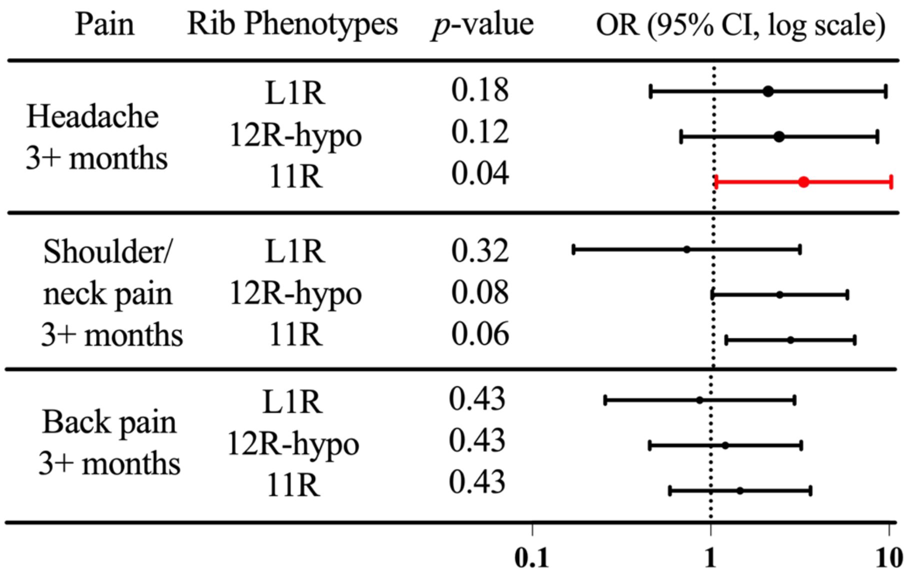

**Fig. S3.**

Associations between variant status and the experience of pain lasting 3+ months (including headaches, neck/shoulder pain, and back pain). 95% confidence intervals of odds ratio (OR) are shown. No test reaches a statistically significant threshold (FDR adjusted *p*-values < 0.05). Abbreviations: Heterozygous = Het; Hemizygous = Hemi. Only *GPC3* has Hemi as it is the only gene on the X chromosome.

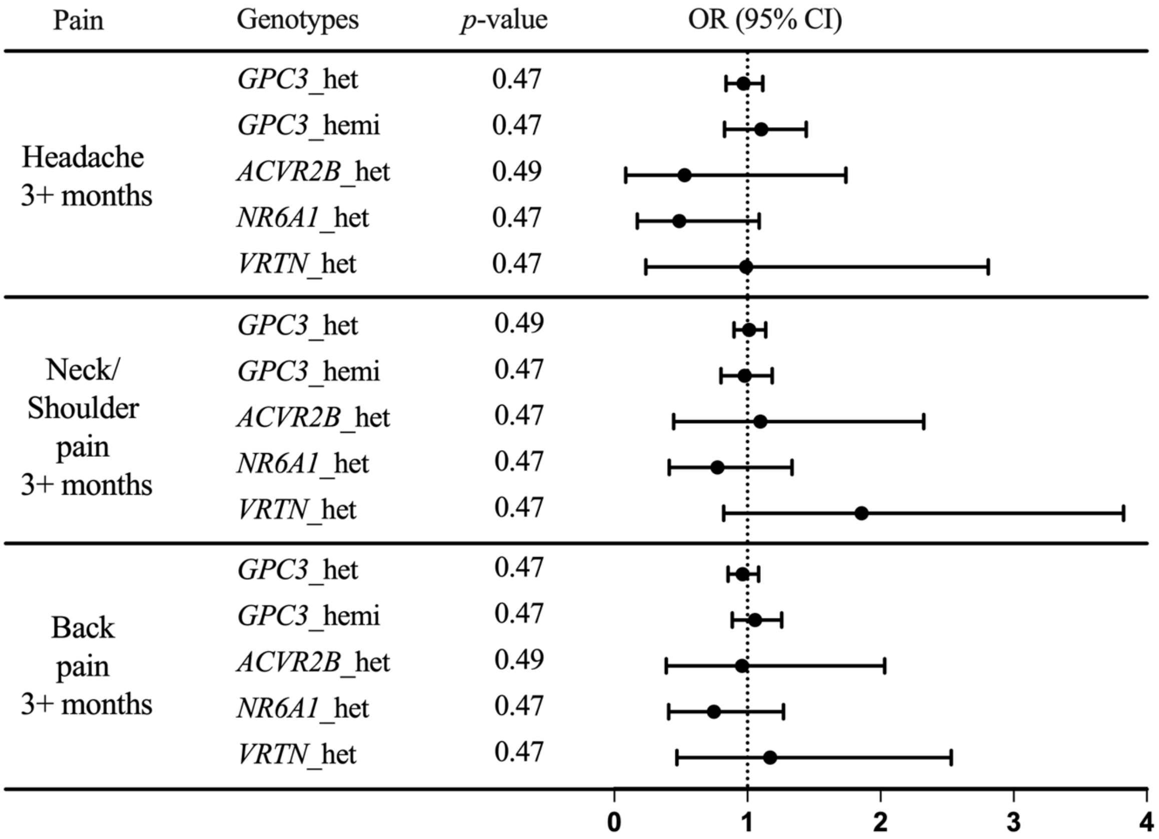

**Fig. S4.**

Association between chronic pain outcomes (headache, neck/shoulder pain, and back pain for 3+ months) and predictor variables, including genetic variant carriers in green (grouped *GPC3*, *VRTN*, *ACVR2B*, and *NR6A1* variants) and vertebral anomalies in orange (grouped six lumbar vertebrae, transitional S1, complete sacralization, and four lumbar vertebrae). One-sided tests were performed, and *p*-values were adjusted using the false discovery rate (FDR) method. Adjusted p-values and odds ratios (OR) with 95% confidence intervals (CI) are reported. A significant association (*FDR-adjusted $p < 0.05$) is observed between vertebral anomalies and chronic headache.

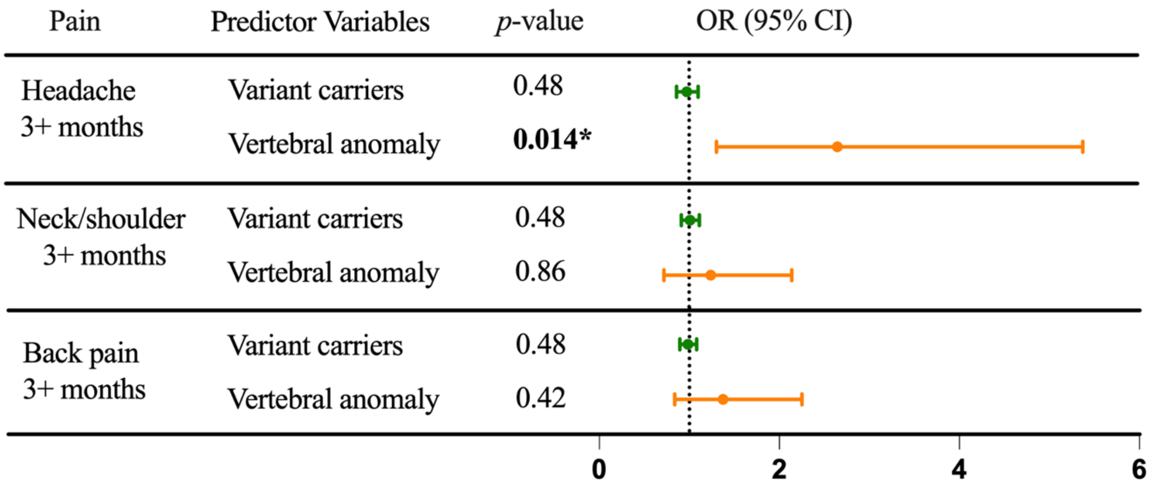

**Fig. S5.**

A schematic representation of the candidate gene and variant selection process. A gene panel was first selected through known clinical or phenotypic associations. (A) Genes collected from the Human Phenotype Ontology (HPO) database. (B) Genes collected from the Mouse Genome Informatics (MGI) phenotype databases, along with additional gene *NR6A1* identified in our previous mouse studies.

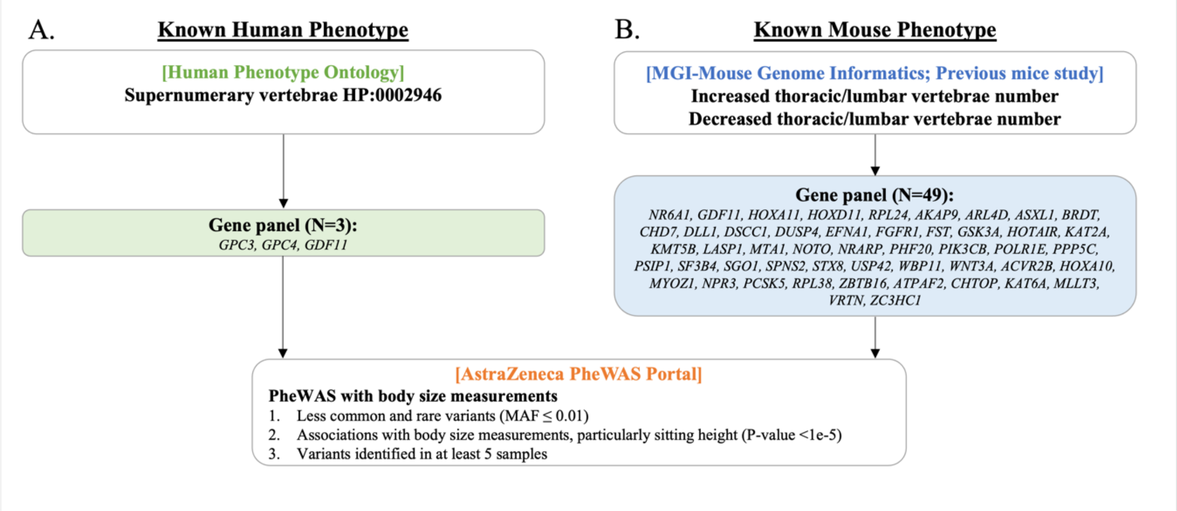

**Fig. S6.**

Flowchart for vertebral and rib phenotyping. (A) When C2 is visible on the body MRI, vertebrae are counted sequentially from C2 down to the last presacral segment. (B) When C2 is not visible on the MRI, alignment between DXA and MR scans is achieved through dense correspondence matching. Intervertebral discs are labelled on the MRI and overlaid onto the whole-body DXA scan. T1 is identified according to the location of the first rib pairs on the DXA images. Vertebral and rib phenotypes are then annotated accordingly. Reproduced by kind permission of UK Biobank ©.

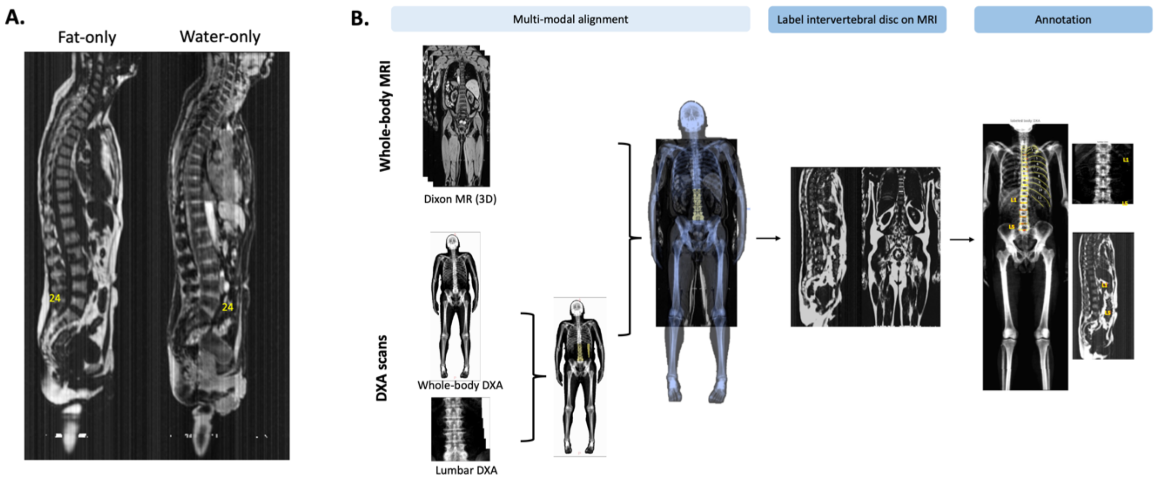

**Fig. S7.**

Example Dixon MR and DXA (total body and lumbar) images illustrating the vertebral and rib phenotypes assessed in this study. (A) Five vertebral phenotypes, presented from the most to the least number of vertebral segments: 6 lumbar vertebrae, lumbarised S1, normal (5 lumbar vertebrae), complete sacralisation, and 4 lumbar vertebrae. Additional or fewer vertebral segments are indicated by red and cyan arrows, respectively. (B) Rib phenotypes, examined based on high-resolution lumbar DXA images, include the presence of extra L1 ribs, 12 rib pairs, hypoplasia of the 12th ribs, and aplasia of the 12th ribs. Additional or fewer ribs compared to the normal 12 rib pairs are indicated by magenta and green arrows, respectively. Reproduced by kind permission of UK Biobank ©.

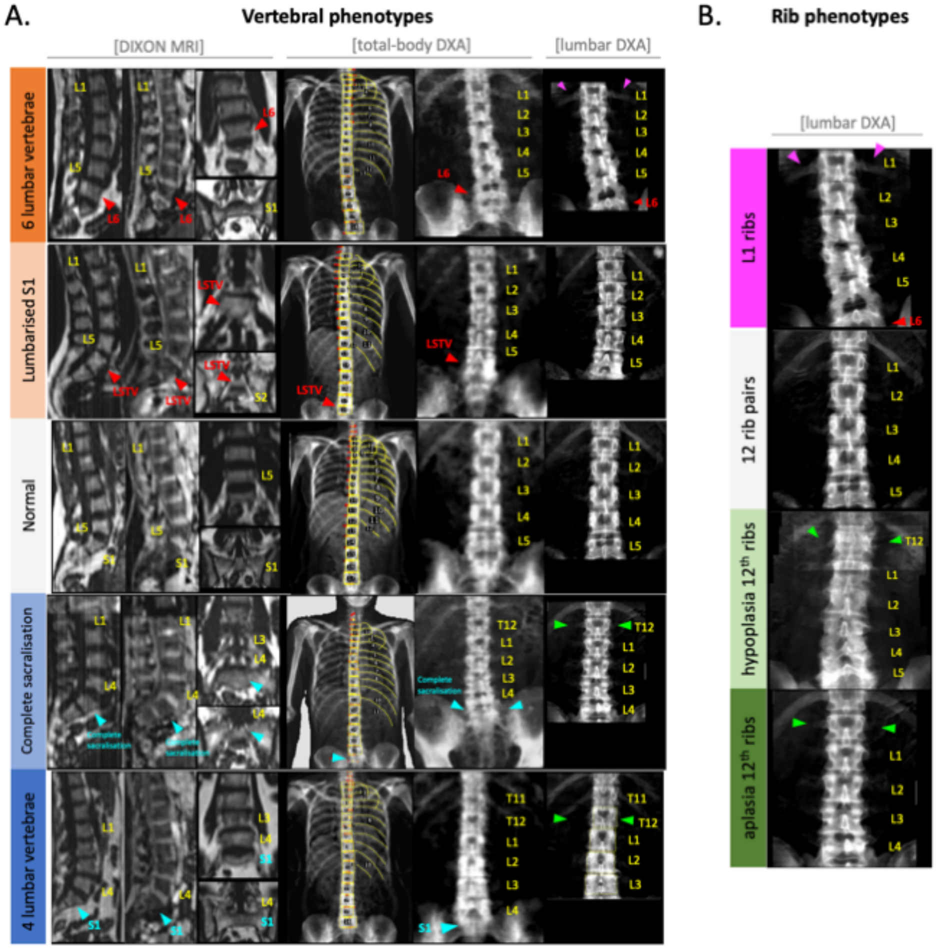

**Table S1.**

Mann-Whitney U test results comparing the distribution of vertebral and rib phenotypes between males and females across different genotypes. The table presents the phenotype type, genotypes, Mann-Whitney U statistic, and p-values for each test. No comparison reaches a statistically significant threshold.

| **Phenotype** | **Genotype** | **Mann-Whitney U Statistic** | ***p-value*** |
|---|---|---|---|
| Vertebral | Non-carriers | 31839.50 | 0.52 |
| Vertebral | *ACVR2B* het | 40.00 | 0.28 |
| Vertebral | *NR6A1* het | 27.50 | 0.09 |
| Vertebral | *VRTN* het | 3.50 | 0.81 |
| Ribs | Non-carriers | 30781.50 | 0.54 |
| Ribs | *ACVR2B* het | 30.00 | 0.28 |
| Ribs | *NR6A1* het | 12.50 | 0.44 |
| Ribs | *VRTN* het | 3.50 | 0.81 |

**Table S2.**

Association between vertebral phenotypes and health-related outcomes. Logistic regression was used to estimate the association between vertebral anomaly categories (reference: normal anatomy) and disease outcomes, with sex as a covariate. Adjusted p-values are reported after two levels of false discovery rate (FDR) correction: (1) within each disease for four vertebral phenotypes, and (2) within each ICD-10 subcategory (i.e. G90-G99: Other disorders of the nervous system). Significant p-values after the second FDR correction are highlighted in bold. Sample sizes for each vertebral phenotype category were as follows: normal anatomy (n = 805), 6 lumbar vertebrae (n = 49), transitional S1 (n = 38), complete sacralisation (n = 35), and 4 lumbar vertebrae (n = 15). *: adjusted P-value<0.05 **: adjusted P-value<0.01.

| | Six lumbar | Transitional S1 | Complete sacralisation | Four lumbar |
|---|---|---|---|---|
| Nervous system disorders | G93 Other disorders of brain | G50 Disorders of trigeminal nerve | | G24 Dystonia |
| Eye and adnexa disorders | **H02 Other disorders of eyelid*** | **H52 Disorders of refraction and accommodation*** | | |
| Ear and mastoid process disorders | | | | H61 Other disorders of external ear |
| Circulatory system disorders | | | | I25 Chronic ischaemic heart disease<br>I87 Other disorders of veins |
| Respiratory system disorders | | | J01 Acute sinusitis<br>J04 Acute laryngitis and tracheitis | |
| Digestive system disorders | | | K64 Haemorrhoids and perianal venous thrombosis | K06 Other disorders of gingiva and edentulous alveolar ridge<br>K22 Other diseases of oesophagus<br>**K44 Diaphragmatic hernia****<br>K59 Other functional intestinal disorders<br>**K92 Other diseases of digestive system*** |
| Musculoskeletal system and connective tissue disorders | M13 Other arthritis | | **M47 Spondylosis*** | M47 Spondylosis |
| Genitourinary system disorders | | N13 Obstructive and reflux uropathy | N34 Urethritis and urethral syndrome | N34 Urethritis and urethral syndrome |

**Table S3.**

**Summary of associations between health-related outcomes and the four variant carrier status.** Logistic regression was used to estimate the association between variant carrier status and disease outcomes. Sex and ancestry are included as covariates. Adjusted p-values are reported after false discovery rate (FDR) correction for the four variants within each ICD-10 subcategory (i.e. G90-G99: Other disorders of the nervous system). *: adjusted p-value<0.05; **: adjusted p-value<0.01.

| **Category** | ***GPC3*** **Carriers** | ***NR6A1*** **Carriers** | ***VRTN*** **Carriers** | ***ACVR2B*** **Carriers** |
|---|---|---|---|---|
| Digestive system disorders | | | K13: other diseases of lip and oral mucosa** | |
| Eye and adnexa disorders | | | | H25: senile cataract* |
| Genitourinary system disorders | | N76: other inflammation of vagina and vulva* | | |
| Musculoskeletal system and connective tissue disorders | | | | M46: other inflammatory spondylopathies** |
| Respiratory system disorders | J40: bronchitis, not specified as acute or chronic* | | J43: emphysema*<br>J45: asthma* | |
